# Diet, physical activity and behavioural disinhibition in middle-aged and older adults: a UK Biobank study

**DOI:** 10.1101/2020.12.04.20243824

**Authors:** Lizanne JS Schweren, Daan van Rooij, Huiqing Shi, Alejandro Arias-Vasquez, Lin Li, Henrik Larsson, Liv Grimstvedt Kvalvik, Jan Haavik, Jan Buitelaar, Catharina A Hartman

**Affiliations:** Interdisciplinary Center Psychopathology and Emotion regulation, University Medical Center Groningen, UMCG HPC CC72. Hanzeplein 1, 9700 RB Groningen, the Netherlands.; Donders Center for Brain, Cognition and Behaviour, Department of Cognitive Neuroscience, RadboudUMC. Geert Grooteplein Zuid 10, 6525 GA Nijmegen, the Netherlands; Donders Center for Brain, Cognition and Behaviour, Department of Cognitive Neuroscience, and Departments of Psychiatry and Human Genetics, RadboudUMC. Geert Grooteplein Zuid 10, 6525 GA Nijmegen, the Netherlands; School of Medical Sciences, Örebro University. Fakultetsgatan 1, 702 81 Örebro, Sweden; School of Medical Sciences, Örebro University. Fakultetsgatan 1, 702 81 Örebro, Sweden. Department of Medical Epidemiology and Biostatistics, Karolinska Institutet. Solnavägen 1, 171 77 Stockholm, Sweden; Department of Biomedicine, University of Bergen. 5007 Bergen, Norway; Department of Biomedicine, University of Bergen. 5007 Bergen, Norway. Bergen Centre of Brain Plasticity, Division of Psychiatry, Haukeland University Hospital. Jonas Lies vei 65, 5021 Bergen, Norway; Donders Center for Brain, Cognition and Behaviour, Department of Cognitive Neuroscience, and Karakter Child and Adolescent Psychiatry University Centre, RadboudUMC. Geert Grooteplein Zuid 10, 6525 GA Nijmegen, the Netherlands; Interdisciplinary Center Psychopathology and Emotion regulation, University Medical Center Groningen. Hanzeplein 1, 9700 RB Groningen, the Netherlands

**Keywords:** Behavioural disinhibition, dietary habits, physical activity, prudent diet, lifestyle, epidemiology

## Abstract

**Background and aims:** Behavioural disinhibition is a prominent feature of multiple psychiatric disorders, and has been associated with poor long-term somatic health outcomes. Modifiable lifestyle factors including diet and moderate-to-vigorous physical activity (MVPA) may be associated with behavioural disinhibition, but their shared and unique contributions have not previously been quantified.

**Methods:** N=157,354 UK Biobank participants who completed the online mental health assessment were included (age 40-69, 2006-2010). Using principal component analyses, we extracted a single disinhibition score and four dietary component scores (prudent diet, elimination of wheat/dairy/eggs, meat consumption, full-cream dairy consumption). In addition, latent profile analysis assigned participants to one of five empirical dietary groups: moderate-healthy, unhealthy, restricted, meat-avoiding, low-fat dairy. Participants self-reported MVPA in minutes/week. Disinhibition was regressed on the four dietary components, the dietary grouping variable and MVPA.

**Results:** in men and women, behavioural disinhibition was negatively associated with prudent diet scores, and positively associated with wheat/dairy/eggs elimination. In men only, disinhibition was associated with consumption of meat and full-cream dairy products. Comparing groups, disinhibition was lower in the moderate-and-prudent diet (reference) group compared to all other groups. Absolute βs ranged from 0.02-0.13 indicating very weak effects. Disinhibition was not associated with MVPA.

**Conclusions:** Among middle-aged and older adults, behavioural disinhibition is associated with multiple features of diet. While the observational nature of UK Biobank does not allow causal inference, our findings foster specific hypotheses (e.g. early malnutrition, elevated immune-response, dietary restraint) to be tested in alternative study designs.

## Background

Behavioural disinhibition is the tendency to act in an uncontrolled fashion, without prior risk-assessment and/or in disregard of social conventions. Manifestations of disinhibition may include poor self-regulation, impulsivity, compulsivity, emotional instability, aggression and high-risk behaviours (1). Extreme manifestations of disinhibition are at the core of severe and often persistent psychiatric disorders, including attention-deficit/hyperactivity disorder (ADHD), mania and addiction. Poor self-regulation and psychiatric disorders related to disinhibition have been associated with a range of negative long-term health outcomes including elevated risks of obesity (2) and cardiovascular disease (3).

Dietary habits may mediate the association between (extreme manifestations of) disinhibition and poor health. Impulsivity and poor self-regulation have been associated with unhealthy food choices, both in experimental settings (4,5) and in observational studies (6). Similarly, overall diet quality is poorer in patients with ADHD (7), mania (8) and addictions (9) compared to unaffected individuals. While a causal pathway running from disinhibition/self-regulation to dietary habits is plausible, dietary intake may also contribute to disinhibited behaviours. Rodents being fed a high-sugar or high-fat diet for a prolonged period of time showed increases in both impulsive (10) and compulsive behaviours (11). Excessive consumption of palatable foods may evoke lasting changes in cortico-limbic dopamine signalling (12). Others have speculated that disinhibited behaviours such as those seen in ADHD might arise as a result of diet-induced inflammation (13) or hypersensitivity to specific foods/allergens (14).

The available literature has several important limitations, however. First, prior studies have primarily focussed on overall diet quality; associations between disinhibition and consumption of specific foods remain largely unexplored. In the current study, we investigated associations between disinhibition and multiple data-driven dietary patterns, and compared homogenous dietary subgroups. Second, behavioural disinhibition is often studied in the young, especially in boys and young men. Dietary patterns differ by sex and change with age (15,16), as might manifestations of disinhibition. Here, we studied disinhibition in the unprecedentedly large population of middle-aged and older UK Biobank participants, allowing investigation of its lifestyle correlates at different ages and for men and women separately. Third, dietary habits inevitably cluster together with other health behaviours including physical activity (17). Lower levels of moderate-to-vigorous physical activity (MVPA) have been associated with poor self-control/disinhibition (18–20), and long-term physical activity interventions have been proposed to ameliorate inhibitory control task performance and reduce symptoms of psychiatric outcomes related to disinhibition (e.g. (21,22)). Prior epidemiological studies, however, focused on either dietary habits or physical activity levels. Here, combined modelling of dietary habits and physical activity allows quantification of their unique and shared contributions.

## Methods

### Study

The data underlying this article were provided by UK Biobank. UK Biobank is a prospective cohort study providing detailed characterisation of over half a million UK-based persons aged 40-69 years at recruitment (2006-2010; http://www.ukbiobank.ac.uk/wp-content/uploads/2011/11/UK-Biobank-Protocol.pdf). With linkage to routinely collected data such as those produced by the National Health Service, UK Biobank offers a highly efficient resource for observational epidemiology (23). UK Biobank has ethical approval from the North West Multi-centre Research Ethics Committee. All participants gave informed consent. For the current study, we included participants who completed the follow-up online mental health questionnaire (MHQ) that was sent to 339,092 participants who had agreed to being contacted by email. Of those, 46% (N=157,354) completed the assessment (24).

### Behavioural disinhibition

To assess disinhibition as a unitary construct, we performed a principal component analysis (PCA) on all disinhibition-related items. For details, see Supporting Information (SI) Section 1. First, all data-fields related to disinhibition, impulsivity, compulsivity and/or emotional instability were selected. Selected data-fields covered addictions including smoking, risk behaviours such as heavy drinking, self-reported and hospital-record diagnoses of selected psychiatric disorders, self-harm behaviours and personality questionnaire items. To ascertain a balanced representation of different manifestations of disinhibition, we defined nine types of disinhibited behaviours (Table 1). Next, we performed a PCA based on tetrachoric correlations between these behaviours. The single-component model, preferred *a priori*, presented with no interpretational shortcomings: all behaviours loaded positively on the principal component with factor loadings ranging from 0.335 to 0.708 (Table 1). For each subject, a factor score was extracted with higher scores indicating a higher tendency for disinhibition.. Extreme values were trimmed at 97.5% and assigned that score, resulting in disinhibition scores ranging from −0.78 to 3.40 (M=-0.06, SD=1.07).

**Table 1:** Cases per behavioural group, its sensitivity and factor loading.

| Group | N (%) | Loading | Definition |
| --- | --- | --- | --- |
| Addiction | 9,534 (6.1) | 0.708 | a) participant self-reports past or current addiction – OR –<br>b) hospital diagnosis of substance dependence syndrome |
| Self-harm | 6,871 (4.4) | 0.637 | a) participant self-reports past or current self-harm |
| Mania | 25,763 (16.4) | 0.613 | a) participant self-reports diagnosis of mania by professional – OR –<br>b) hospital diagnosis of mania/bipolar disorder – OR –<br>c) participant has experienced period with at least one symptom of mania |
| OCD | 996 (0.6) | 0.591 | a) participant self-reports diagnosis of OCD by professional – OR –<br>b) hospital diagnosis of OCD/tic disorder/trichotillomania/anankastic PD |
| Externalising | 134 (0.1) | 0.589 | a) participant self-reports diagnosis of ADHD by professional – OR –<br>b) hospital diagnosis of any externalising disorder |
| Cannabis use | 11,205 (7.1) | 0.552 | a) participant self-reports having used cannabis >10 times in life |
| Personality | 23,892 (15.2) | 0.536 | a<sub>1</sub>) participant self-reports moods going up and down – AND –<br>a<sub>2</sub>) participant self-reports being easily fed-up – AND –<br>a<sub>3</sub>) participant self-reports being irritable – OR –<br>b) hospital diagnosis of emotionally unstable PD |
| Smoking | 7,615 (4.8) | 0.475 | a) participant self-reports current daily smoking |
| Risk-taking | 23,994 (15.3) | 0.335 | a) participant self-reports weekly binge-drinking – OR –<br>b) hospital diagnosis of acute intoxication with alcohol/drugs |

Abbreviations: ADHD = attention-deficit/hyperactivity disorder; OCD = obsessive-compulsive disorder; PD = personality disorder

### Diet

Dietary habits were assessed using a 29-item food frequency questionnaire (FFQ) about participants’ past-year average diets. Individual items are highly correlated reflecting underlying dietary patterns (25). We thus performed two established data-reduction techniques: PCA to detect shared variance between items (i.e. dietary components), and latent profile analysis to detect clusters of subjects behaving similarly with respect to the dietary components (i.e. dietary groups). For details see SI Section 2 and 3.

Raw data included continuous variables (e.g. tablespoons of vegetables), ordinal variables (e.g. cereal intake frequency), food type descriptions (e.g. bread type: brown/white/wholemeal/other), and one tick-box item assessing elimination of specific food groups (eggs/dairy/wheat/sugar). Alcohol-related items were removed, since heavy drinking contributed to disinhibition scores. Binary contrasts were created for each food type (e.g. ‘brown bread’ vs. ‘any other bread type’) and elimination item (e.g. ‘I never eat eggs’ vs. ‘no restrictions’). The resulting 58 food items/contrasts, regressed on age and sex, were entered in a PCA with promax rotation on the residuals, starting with one component and adding components one-by-one. Components were retained as long as they remained unique, interpretable and stable. The optimal outcome contained four dietary components (Table 2). DC1 reflects a prudent diet, with positive loadings for fruits, vegetables, wholegrain bread and fish, and negative loadings for refined carbohydrate products, processed meat and instant coffee. DC2 reflects elimination of wheat, dairy and/or eggs. DC3 reflects meat and to a lesser extent fish consumption, and DC4 reflects consumption of full-cream milk and butter/spreads. Correlations between components were modest (r≤0.124)

**Table 2.**
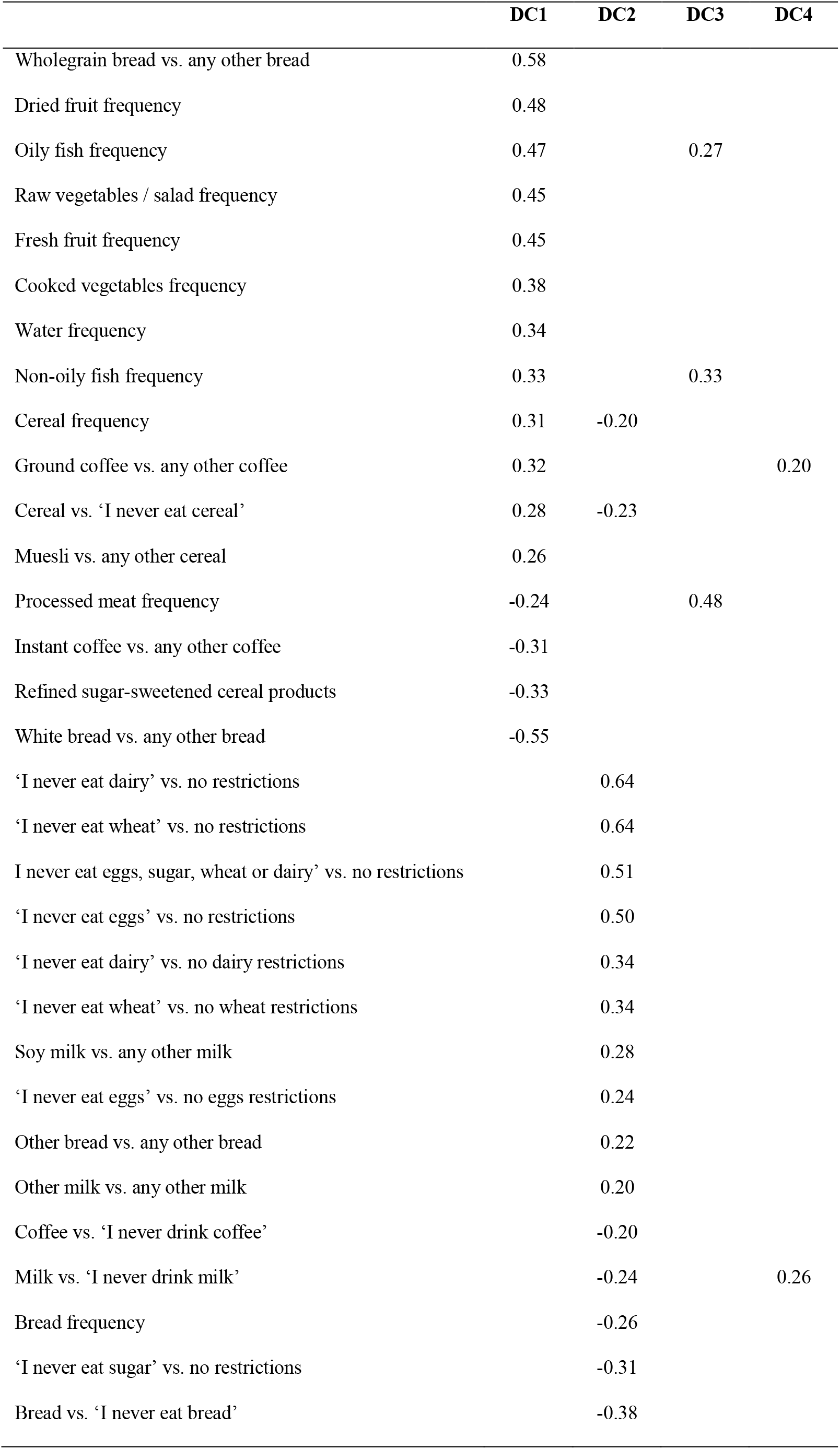
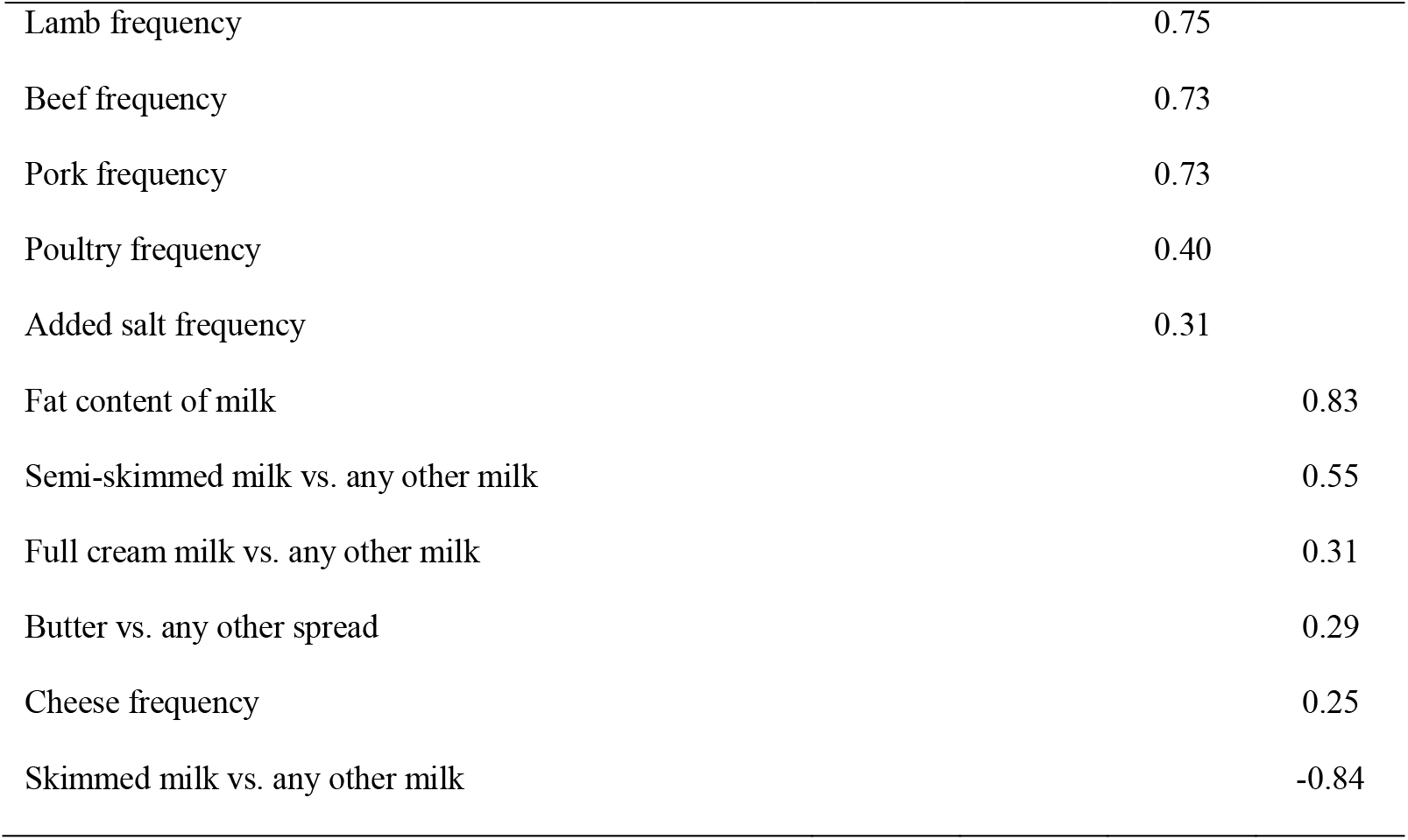
Factor loadings (≤-0.20 or ≥0.20) per dietary component (DC1-4)

Next, we performed latent profile analysis (R-package ‘mclust’ (26)) based on the four diet scores to derive relatively homogenous participant groups/clusters. Based on model fit, parsimony, stability, group size and plausibility, the optimal model comprised five mutually exclusive latent groups (Table 3, Figure 1, SI Section 3). The largest group (n=49,463, 31.4%) had relatively high prudent diet scores and moderate scores across the other three DCs and was set as the reference group (‘moderate-and-prudent’). The second group was characterised by a generally unhealthy (i.e. non-prudent) diet and high consumption of full-cream dairy products (‘unhealthy’, n=42,663, 27.1%). The third group comprised individuals who avoided meat and adhered to a prudent diet (‘avoid meat’, n=21,797, 13.9%). The remaining two groups were each driven by a single DC: one group comprised participants who avoided full-cream dairy products (‘low-fat dairy’: n=32,555, 20.7%), and one group comprised participants who eliminated wheat/dairy/eggs from their diet (‘restricted’; n=10,876, 6.9%).

**Table 3.** Descriptive statistics per dietary group

|  | **Moderate-and-prudent** | **Unhealthy** | **Low-fat dairy** | **Avoid meat** | **Restricted** |
| --- | --- | --- | --- | --- | --- |
|  | **M(SD) / N(%)** | **M(SD) / N(%)** | **M(SD) / N(%)** | **M(SD) / N(%)** | **M(SD) / N(%)** |
| N | 49,463 (31.4) | 42,663 (27.1) | 32,555 (20.7) | 21,797 (13.9) | 10,876 (6.9) |
| Male | 20,315 (41.1) | 20,143 (47.2)<sup>a</sup> | 12,054 (37.0)<sup>a</sup> | 11,287 (51.8)<sup>a</sup> | 4,461 (41.0)<sup>a</sup> |
| Age | 55.5 (7.7) | 55.9 (7.8)<sup>bc</sup> | 56.3 (7.6)<sup>bc</sup> | 56.2 (7.9)<sup>ab</sup> | 56.4(7.7)<sup>bc</sup> |
| BMI | 26.2 (4.3) | 27.4 (4.7)<sup>bc</sup> | 27.3 (4.7)<sup>bc</sup> | 26.1 (4.3)<sup>c</sup> | 26.7 (4.8)<sup>bc</sup> |
| IMD | 13.4 (10.7) | 15.7 (12.6)<sup>bc</sup> | 14.5 (11.7)<sup>bc</sup> | 15.5 (12.1)<sup>bc</sup> | 16.6 (13.2)<sup>bc</sup> |
| Years in education | 17.1 (4.1) | 15.6 (4.7)<sup>bc</sup> | 16.3 (4.5)<sup>bc</sup> | 17.0 (4.3)<sup>bc</sup> | 16.0 (4.7)<sup>bc</sup> |
| Annual income<sup>d</sup> | 32.9 (16.0) | 29.1 (14.9)<sup>bc</sup> | 31.8 (16.0)<sup>bc</sup> | 32.2 (16.7)<sup>b</sup> | 30.6 (16.4)<sup>bc</sup> |
| Unemployment | 524 (1.1) | 672 (1.6)<sup>b</sup> | 394 (1.2) | 289 (1.3) | 177 (1.6)<sup>b</sup> |
| Non-white ethnicity | 956 (1.9) | 1,136 (2.7)<sup>bc</sup> | 697 (2.1)<sup>b</sup> | 1,209 (5.6)<sup>bc</sup> | 561 (5.2)<sup>bc</sup> |
| Sleep duration |  |  |  |  |  |
| Short | 9,592 (19.4) | 9,762 (22.9)<sup>bc</sup> | 7,271 (22.3)<sup>bc</sup> | 4,862 (22.3)<sup>bc</sup> | 2,831 (26.0)<sup>bc</sup> |
| Long | 782 (1.6) | 1,111 (2.6)<sup>bc</sup> | 721 (2.2)<sup>bc</sup> | 527 (2.4)<sup>bc</sup> | 326 (3.0)<sup>bc</sup> |
| DC1 (prudent diet) | 0.55 (0.62) | -0.92 (0.72) | 0.10 (0.95) | 0.42 (0.92) | -0.04 (1.12) |
| DC2 (no wheat/dairy/eggs) | -0.46 (0.37) | -0.25 (0.38) | -0.15 (0.49) | 0.36 (0.56) | 2.84 (1.54) |
| DC3 (meat) | 0.14 (0.78) | 0.34 (0.77) | 0.05 (0.94) | -0.90 (1.21) | -0.29 (1.26) |
| DC4 (full-cream dairy) | 0.53 (0.51) | 0.45 (0.47) | -1.55 (0.39) | 0.44 (0.69) | -0.38 (1.10) |
| MVPA quintiles | 3.06 (1.36) | 2.82 (1.43)<sup>bc</sup> | 2.96 (1.39)<sup>bc</sup> | 3.11 (1.40)<sup>bc</sup> | 3.03 (1.43) |

a. significantly different from the moderate-and-healthy diet group in unadjusted analyses; b. significantly different from the moderate-and-prudent diet group in unadjusted analyses for men; c. significantly different from the moderate-and-prudent diet group in unadjusted analyses for women. d. \*1,000 euros, adjusted for household size.

**Figure 1.**
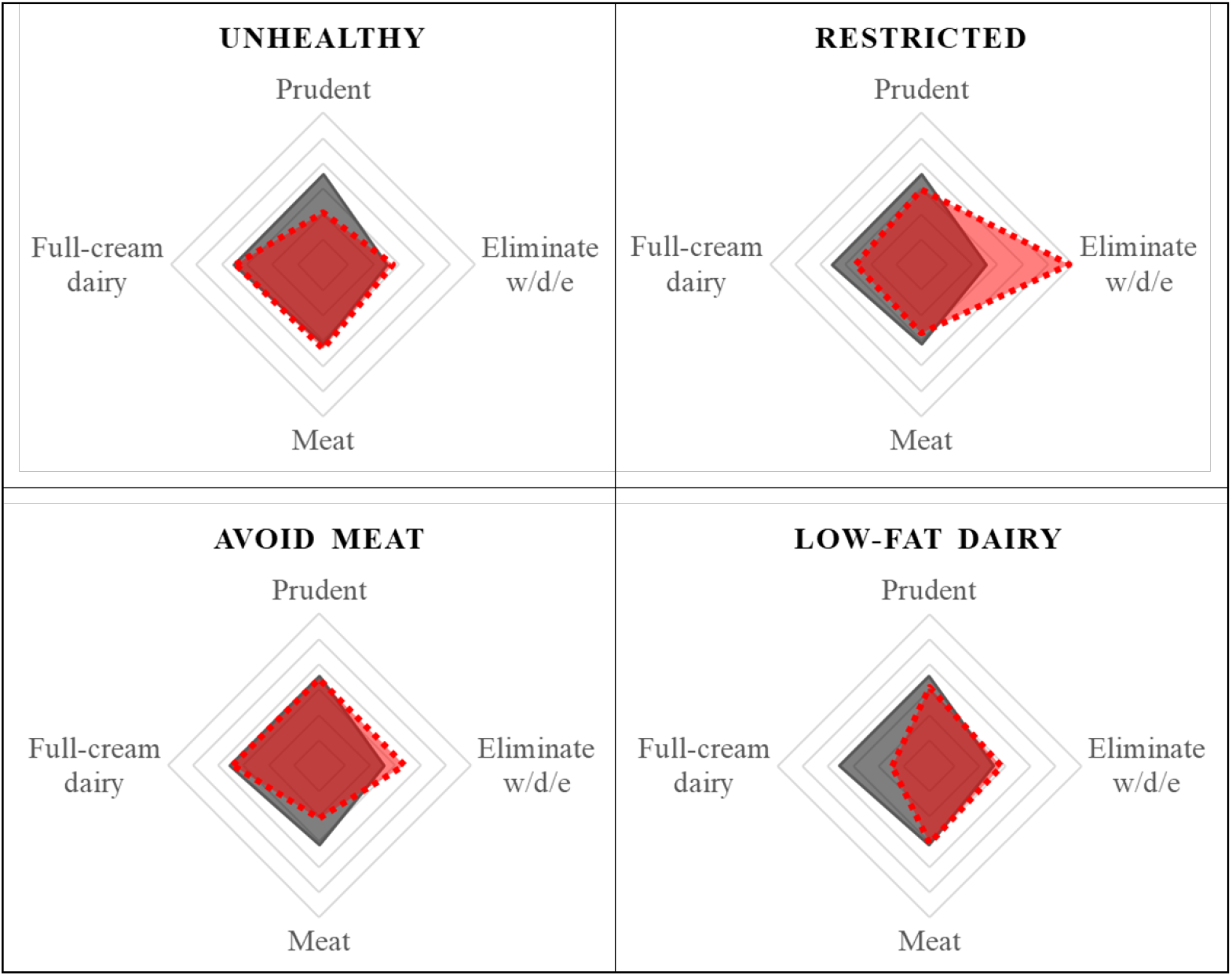
Average scores on dietary components DC1-4 of each data-driven dietary group (in red/dashed), projected over the dietary profile of the moderate-and-prudent diet group (in grey/solid). Abbreviations: w/d/e = wheat/dairy/eggs.

### Physical activity

Participants self-reported their frequency (days/week) and duration (minutes/day) of engaging in moderate and vigorous physical activity. MVPA was calculated as the sum of moderate and vigorous physical activity in minutes/week. MVPA was strongly skewed, ranging from 0 to 8,640 minutes/week with a median of 180 minutes/week and 80% of participants being active for ≤480 minutes/week. MVPA was thus converted to pseudo-continuous quintile scores: 1: 0-43 minutes/week; 2: 43-120 minutes/week; 3: 120-240 minutes/week; 4: 240-480 minutes/week; 5: >480 minutes/week.

In sensitivity analyses, we also evaluated moderate and vigorous physical activity separately. For MPA, quintile scores were created as before (1: 0-30 minutes/week, 2: 30-80 minutes/week, 3: 80-151 minutes/week, 4: 151-360 minutes/week, 5: >360 minutes/week). A large proportion of the sample never performed VPA, thus VPA was split in three groups (1: never; 2: 0-median i.e. 0-80 minutes/week; 3: >80 minutes/week).

### Statistical procedures

The following covariates were included in each model: ethnicity (white/other), body mass index in kg/m^2^, self-reported habitual sleep duration (short: <7 hours/night; normal: 7-9 hours/night or 7-8 hours/night for adults age ≥65; long: >9 *resp*. >8 hours/night), neighbourhood-based index of multiple deprivation (IMD) by region (England, Scotland, Wales), self-reported annual household income adjusted for household size, self-reported unemployment status (unemployed/other), and years of education (27) (no qualifications: 7y; CSEs: 10y; O levels/GCSEs:10y; A levels/AS levels:13y; other professional qualification: 15y; NVQ or HNC: 19y; college or university degree: 20y). Missing values for all variables were imputed with multivariate imputation using chained equations (R-package ‘MICE’ (28)).

Subsequent steps were performed in men and women separately. All variables were standardised. For descriptive purposes, we first predicted disinhibition from all covariates. Next, in six single-predictor models, we predicted disinhibition from DC1-4, the dietary grouping variable (reference group=‘moderate-and-prudent’) and MVPA. Initially, an age-by-predictor interaction term was included in each model as well; however, no relevant interactions were detected (SI Section 4), hence age-interaction terms were discarded. To assess unique associations between disinhibition and each predictor, we ran two multiple-predictor models: one predicting disinhibition from the DC1-4 and MVPA, and one predicting disinhibition from the dietary grouping variable and MVPA. Again, age-interactions were discarded upon finding no relevant interaction effects.

Alpha was conservatively adjusted for testing four DCs, one dietary grouping variable with five levels, and MVPA, in men and women separately: α=0.05/((4+(5-1)+1)*2)=0.0028. Provided power=0.8, α=0.0028 and n>66,419, we were able to detect associations of β≥0.02. Differentiation between statistically and clinically significant findings is not achieved by further lowering of α, but by investigating β. Absolute βs above 0.2 are generally referred to as ‘weak’; however, in models predicting complex behaviours, large βs are not expected and small βs can be informative (29). Consensus regarding denotation is lacking. Here, for interpretational purposes, we qualify absolute βs of significant associations (p<0.0028) between 0.02-0.19 as very weak, 0.2-0.39 as weak, 0.4-0.59 as moderate, 0.6-0.79 as strong and ≥0.8 as very strong. Absolute β<0.02 is qualified as not associated.

## Results

### Disinhibition

Disinhibition scores were higher in men (M=0.01, SD=1.09) compared to in women (M=-0.11, SD=1.05; β_FEMALE_=-0.118, se=0.005, p<0.0001). In the covariate-only model, disinhibition was associated with younger age (men: β=-0.226, se=0.004; women: β=-0.212, se=0.003), unemployment (men: β=0.215, se=0.027; women: β=0.275, se=0.035), white ethnicity (men: β_NON-WHITE_=-0.189, se=0.022; women: β_NON-WHITE_=-0.123, se=0.019), long sleep duration (men: β=0.201, se=0.024; women: β=0.215, se=0.023), short sleep duration (men: β=0.129, se=0.009; women: β=0.122 se=0.008), neighbourhood deprivation (men: β=0.112, se=0.004; women: β=0.111, se=0.004), and BMI (men: β=0.040, se=0.004; women: β=0.036, se=0.003). In women, disinhibition was also associated with lower adjusted income (β=-0.036, se=0.003) and more years of education (β=0.022, se=0.003).

### Dietary components

In the single-predictor models including all covariates, prudent diet (DC1) was very weakly associated with lower disinhibition scores in men (β=-0.036) and women (β=-0.043; Table 4). By contrast, elimination of wheat, dairy and/or eggs (DC2) was very weakly associated with higher disinhibition scores in both groups (β_MEN_=0.030, β_WOMEN_=0.038). Meat consumption (DC3) and full-cream dairy consumption (DC4) were very weakly associated with more disinhibition in men (β_DC3_=0.041; β_DC4_=0.023) but not in women. Associations between disinhibition and each DC remained virtually unchanged in the multiple-predictor model that included all dietary components simultaneously as well as physical activity scores (Table 4), suggesting a) minimal overlap between the four dietary components, and b) that associations between disinhibition and diet were not accounted for by physical activity.

**Table 4.**
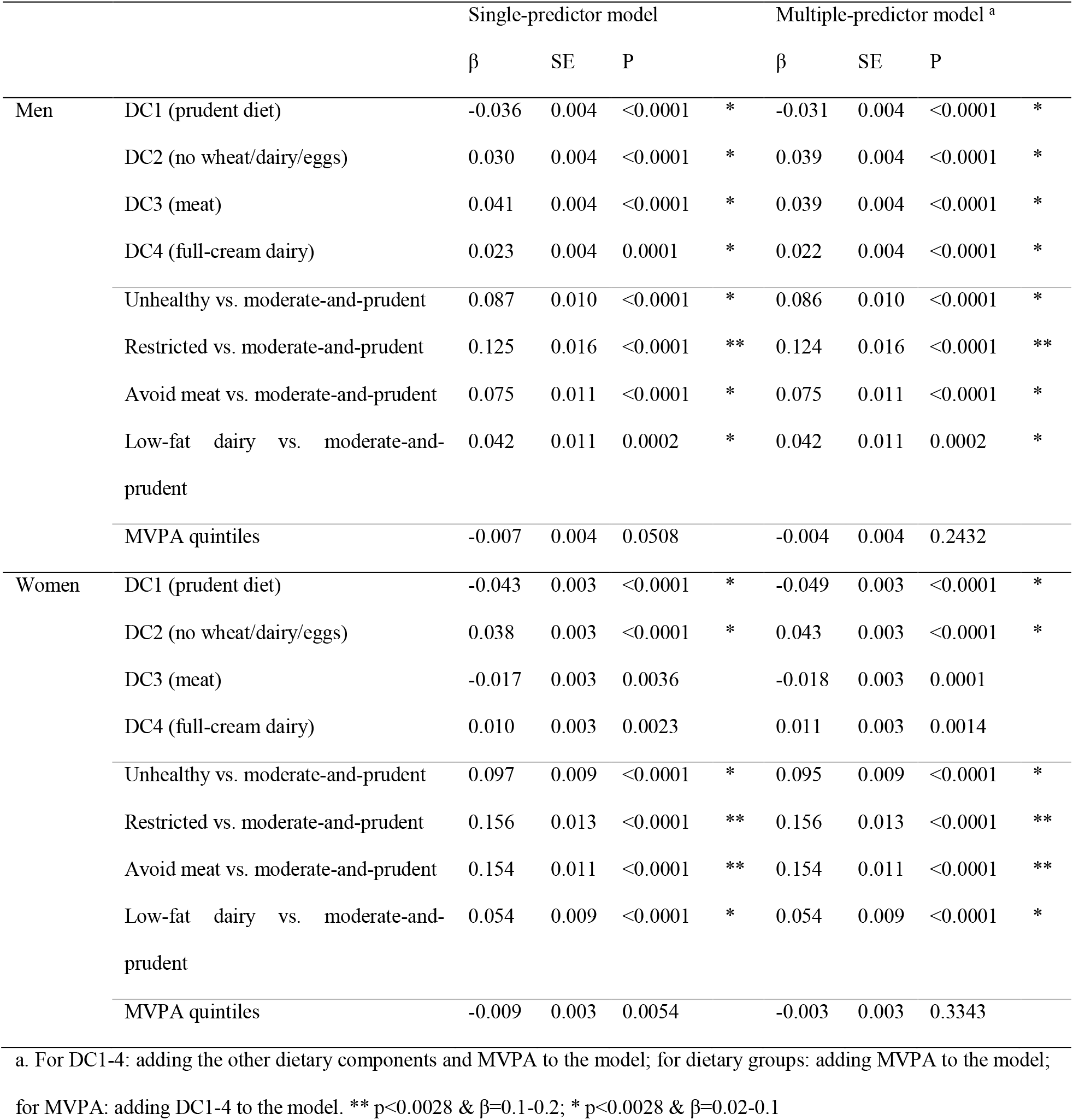
Outcomes

|  |  | Single-predictor model |  |  |  | Multiple-predictor model <sup>a</sup> |  |  |  |
| --- | --- | --- | --- | --- | --- | --- | --- | --- | --- |
| | | $\beta$ | SE | P | | $\beta$ | SE | P | |
| Men | DC1 (prudent diet) | -0.036 | 0.004 | <0.0001 | * | -0.031 | 0.004 | <0.0001 | * |
|  | DC2 (no wheat/dairy/eggs) | 0.030 | 0.004 | <0.0001 | * | 0.039 | 0.004 | <0.0001 | * |
|  | DC3 (meat) | 0.041 | 0.004 | <0.0001 | * | 0.039 | 0.004 | <0.0001 | * |
|  | DC4 (full-cream dairy) | 0.023 | 0.004 | 0.0001 | * | 0.022 | 0.004 | <0.0001 | * |
|  | Unhealthy vs. moderate-and-prudent | 0.087 | 0.010 | <0.0001 | * | 0.086 | 0.010 | <0.0001 | * |
|  | Restricted vs. moderate-and-prudent | 0.125 | 0.016 | <0.0001 | ** | 0.124 | 0.016 | <0.0001 | ** |
|  | Avoid meat vs. moderate-and-prudent | 0.075 | 0.011 | <0.0001 | * | 0.075 | 0.011 | <0.0001 | * |
|  | Low-fat dairy vs. moderate-and-prudent | 0.042 | 0.011 | 0.0002 | * | 0.042 | 0.011 | 0.0002 | * |
|  | MVPA quintiles | -0.007 | 0.004 | 0.0508 |  | -0.004 | 0.004 | 0.2432 |  |
| Women | DC1 (prudent diet) | -0.043 | 0.003 | <0.0001 | * | -0.049 | 0.003 | <0.0001 | * |
|  | DC2 (no wheat/dairy/eggs) | 0.038 | 0.003 | <0.0001 | * | 0.043 | 0.003 | <0.0001 | * |
|  | DC3 (meat) | -0.017 | 0.003 | 0.0036 |  | -0.018 | 0.003 | 0.0001 |  |
|  | DC4 (full-cream dairy) | 0.010 | 0.003 | 0.0023 |  | 0.011 | 0.003 | 0.0014 |  |
|  | Unhealthy vs. moderate-and-prudent | 0.097 | 0.009 | <0.0001 | * | 0.095 | 0.009 | <0.0001 | * |
|  | Restricted vs. moderate-and-prudent | 0.156 | 0.013 | <0.0001 | ** | 0.156 | 0.013 | <0.0001 | ** |
|  | Avoid meat vs. moderate-and-prudent | 0.154 | 0.011 | <0.0001 | ** | 0.154 | 0.011 | <0.0001 | ** |
|  | Low-fat dairy vs. moderate-and-prudent | 0.054 | 0.009 | <0.0001 | * | 0.054 | 0.009 | <0.0001 | * |
|  | MVPA quintiles | -0.009 | 0.003 | 0.0054 |  | -0.003 | 0.003 | 0.3343 |  |

a. For DC1-4: adding the other dietary components and MVPA to the model; for dietary groups: adding MVPA to the model; for MVPA: adding DC1-4 to the model. \*\* $p < 0.0028$ & $\beta = 0.1-0.2$ ; \* $p < 0.0028$ & $\beta = 0.02-0.1$

### Dietary groups

Demographic/socioeconomic differences between the five dietary groups are shown in Table 3. Taking into account all covariates, disinhibition was significantly lower in the moderate-and-prudent diet group compared to all other diet groups (Table 4). In men, the difference in disinhibition compared to the moderate-and-prudent diet group was strongest in the restricted group (β=0.125), weaker in the unhealthy (β=0.087) and meat-avoiding (β=0.075) groups, and least pronounced in the low-fat dairy group (β=0.042). In women, relatively strong effects were found in the restricted diet group (β=0.156) and the meat-avoiding group (β=0.154), followed by the unhealthy group (β=0.097) and the low-fat dairy group (β=0.054). Note, however, that all effects were qualified as very weak (β<0.2). Model estimates were minimally affected by adding MVPA to the model (Table 4).

### Moderate-to-vigorous physical activity

MVPA was not associated with disinhibition in men (β=-0.007) or women (β=-0.009; Table 4). Estimates were further attenuated after adding DC1-4 or the dietary grouping variable to the model. In sensitivity analyses, the models were rerun for moderate (MPA) and vigorous (VPA) physical activity separately. In the single-predictor models, MPA was not associated with disinhibition in men (β=<-0.001; SE=0.004, p=0.9186) or women (β=-0.004; SE=0.003, p=0.1647). VPA was very weakly associated with lower disinhibition in both groups (men: β=-0.023; SE=0.004, p<0.0001; women: β=-0.023; SE=0.003, p<0.0001). Adding the dietary grouping variable to the model, the association between VPA and disinhibition remained significant (men: β=-0.021; SE=0.004, p<0.0001; women: β=-0.021; SE=0.003, p<0.0001), while the association fell short of significance after adding DC1-4 (men: β=-0.018; SE=0.004, p<0.0001; women: β=-0.016; SE=0.003, p<0.0001).

## Discussion

We investigated dietary habits, physical activity and behavioural disinhibition in individuals age 40 and older. Among men and women and across the age range, adherence to a prudent diet was associated with lower disinhibition scores, while elimination of wheat, dairy and/or eggs was associated with higher disinhibition scores. In men, disinhibition was also associated with higher habitual consumption of meat and high-fat dairy products. Classifying participants based on their multivariate diet patterns, disinhibition scores were higher in the unhealthy diet, restricted diet, meat-avoiding and low-fat dairy groups compared to the moderate-and-prudent diet reference group. All associations between diet and disinhibition were very weak (β<0.2). MVPA was not associated with disinhibition.

### Observed associations are weak in strength

Our finding of a negative association between prudent diet and disinhibition among middle-aged and older men and women corroborates and extends the existing disinhibition literature that tends to focus on the young, especially young males. We add to the available knowledge by qualifying the association between prudent diet and disinhibition as ‘very weak’. Short sleep duration, for instance, was >2.5 times more strongly associated with disinhibition, and age and unemployment were >5 times more strongly associated. The weak association strength of this and all other associations detected in our study should be kept in mind when reading the below discussion.

### Early-life malnutrition, food intolerances and dietary restraint

Unexpectedly, the strongest lifestyle predictor of disinhibition was a diet restricted in wheat, dairy and/or eggs. Within the restricted diet group, most individuals eliminated a single food group and each food group was equally often eliminated (wheat: 30.5%; dairy: 27.4%; eggs: 27.6%), suggesting that disinhibition is likely not associated with a lack of nutrients specifically provided by either product. Note that the restricted diet group was also characterised by lower overall diet quality compared to the moderate-and-prudent diet group, potentially suggesting a more general lack of nutrients or residual confounding. Participants in the restricted diet group were more often born outside the UK and of lower socioeconomic status, potentially pointing towards early life exposure to malnutrition. In animals, malnutrition early in life is known to cause impulsiveness in adulthood (30). Alternatively, elimination of specific foods may suggest a higher prevalence of (allergic) food intolerances among impulsive individuals. Common genetic variants have been identified between allergic diseases and psychiatric disorders including ADHD, OCD and addiction (31). Studies of dieting/weight control provide another alternative explanation: highly-impulsive individuals tend to score higher on dietary restraint, a cognitive concept characterised by the intention to limit food intake, among others (32). However, participants were instructed to report dietary habits, not intentions. Finally, higher endorsement of elimination items (“Which of the following do you *never* eat?”) may reflect a higher tendency among high-impulsive individuals compared to low-impulsive individuals to endorse these items, even if they do not fully meet the never-criterion.

Our findings regarding meat consumption were contradictory. In prior studies, meat consumption has been associated with emotional instability and impulsive/thrill-seeking traits (33,34), but the opposite (6,35) and null findings (36,37) have also been reported. Here, in men only, meat consumption was associated with higher disinhibition scores. By contrast, both men and women in the meat-avoiding group had higher disinhibition scores compared to those in the moderate-and-prudent diet group. Higher disinhibition in the meat-avoiding group cannot be not explained by lower intake of omega-3 fatty acids (essential fish oil nutrients with potential limited efficacy in treating symptoms of ADHD (38)): oily fish consumption in the moderate-and-prudent and the meat-avoiding groups were similar (data not shown). Compared to the moderate-and-prudent diet group, the meat-avoiding group had higher levels of wheat/dairy/eggs restrictions, and, similar to the dietary restrictions group, a relatively large proportion of individuals of non-white ethnicity and born outside the UK. The early-malnutrition and food-intolerance hypotheses may thus apply to the meat-avoiding group as well. Alternatively, the decision to (partially) eliminate meat from one’s diet might be influenced by fluctuating societal trends, to which impulsive individuals may be more sensitive.

### No unique association between disinhibition and physical activity

Finally, the absence of an association between MVPA and disinhibition is of note. In line with cardiovascular health benefits being greater for vigorous compared to moderate intensity activities (39), we found a very weak association between disinhibition and VPA but not MPA; however this association was partially accounted for by dietary factors. Similarly, leisure time MVPA may be associated with better outcomes compared to occupational MVPA ((40)), but our data did not allow stratification by MVPA-type. Thorough investigation of specific physical activity characteristics is beyond the scope of the current paper, and is recommended for future studies.

### Strengths and limitations

Strengths of the current study include the application of state-of-the-art data-reduction techniques across modalities to capture complex latent variables. The so-created disinhibition scale robustly showed the expected associations with sociodemographic variables. Associations between diet and disinhibition were assessed both at the food-group level and at the level of multivariate dietary patterns, ensuring ecological validity, and were adjusted for other lifestyle parameters. Finally, socioeconomic status was modelled at the societal-, household- and individual level, lowering the risk of residual confounding. The current study has limitations as well. The cross-sectional and observational nature of our study precludes causal inferences. Observed associations might reflect an effect of disinhibition on dietary choices, an effect of dietary intake on disinhibited behaviours, or both. Another inherent vulnerability of observational studies is the possibility of unmeasured confounding, i.e. both disinhibition and dietary behaviours might be driven by an unmeasured third variable (e.g. genetic confounding). Finally, dietary habits and physical activity levels were self-reported and might be subject to systematic bias (e.g. less accurate reporting in high-impulsive participants).

## Conclusion

We found significant and age-independent associations between dietary habits and behavioural disinhibition among middle-aged and older men and women. Although all associations were of very weak strength, our findings generate specific hypotheses that might be tested in alternative study designs.

## Data Availability

UK Biobank data is available upon application

https://www.ukbiobank.ac.uk/

## Funding statement

This work was supported by the European Union’s Horizon 2020 Research and Innovation Program under grant agreement No 728018. The funding source has had no involvement in the study design, data collection, interpretation of the findings, or writing of this manuscript.

## Conflict of interest statement

JH declares that he has in the past 3 years been a paid speaker for Medice, Biocodex, Shire, and Takeda, all unrelated to the work presented in the submitted manuscript. JB has been in the past 3 years a consultant to / member of advisory board of / and/or speaker for Takeda/Shire, Roche, Medice, Angelini, Janssen, and Servier. He is not an employee of any of these companies, and not a stock shareholder of any of these companies. He has no other financial or material support, including expert testimony, patents, royalties. HL has served as a speaker for Evolan Pharma and Shire/Takeda and has received research grants from Shire/Takeda; all outside the submitted work. The companies were in no way involved in the study design or analyses performed in the submitted manuscript, nor in the reporting of results or interpretation of findings. LJSS, DvR, HS, AA, LL, LGK and CAH declare that they have no affiliations with or involvement in any organization or entity with any financial interest or non-financial interest in the subject matter or materials discussed in this manuscript.

## Author contributions

LJSS: conceptualization, data curation, formal analysis, writing – original draft, writing – review and editing, visualization; DvR: conceptualization, data curation, writing – review and editing; HS: writing – review and editing; AAV: funding acquisition, writing – review and editing; LL: writing – review and editing; HL: supervision, writing – review and editing; LGK: writing – review and editing; JH: supervision, writing – review and editing; JB: conceptualization, supervision, writing – review and editing; CAH: conceptualization, supervision, writing – review and editing.

## Notes

### Funding Statement

This work was supported by the European Union Horizon 2020 Research and Innovation Program under grant agreement No 728018. The funding source has had no involvement in the study design, data collection, interpretation of the findings, or writing of this manuscript.

### Author Declarations

Access to the data was granted by UK Biobank following registration with their system and approval of our research project (project number 23668). The North West Multi-centre Research Ethics Committee oversees ethical approval of UK Biobank research procedures. See https://www.ukbiobank.ac.uk/media/0xsbmfmw/egf.pdf

## References

1. Sharma L, Markon KE, Clark LA. Toward a theory of distinct types of “impulsive behaviors”: a meta-analysis of self-report and behavioral measures. Psychol Bull. 2014;140(2):374–408.

2. Cortese S, Moreira-Maia CR, St Fleur D, Morcillo-Peñalver C, Rohde LA, Faraone S V. Association between ADHD and obesity: A systematic review and meta-analysis. Am J Psychiatry. 2016;173(1):34–43.

3. Kubzansky LD, Park N, Peterson C, Vokonas P, Sparrow D. Healthy Psychological Functioning and Incident Coronary Heart Disease. Arch Gen Psychiatry. 2011;68(4):400.

4. Van Blyderveen S, Lafrance A, Emond M, Kosmerly S, O’Connor M, Chang F. Personality differences in the susceptibility to stress-eating: The influence of emotional control and impulsivity. Eat Behav [Internet]. 2016;23:76–81. Available from: http://dx.doi.org/10.1016/j.eatbeh.2016.07.009

5. Guerrieri R, Nederkoorn C, Jansen A. How impulsiveness and variety influence food intake in a sample of healthy women. Appetite. 2007;48(1):119–22.

6. Bénard M, Bellisle F, Kesse-Guyot E, Julia C, Andreeva VA, Etilé F, et al. Impulsivity is associated with food intake, snacking, and eating disorders in a general population. Am J Clin Nutr. 2019;109(1):117–26.

7. Loewen OK, Maximova K, Ekwaru JP, Asbridge M, Ohinmaa A, Veugelers PJ. Adherence to life-style recommendations and Attention-Deficit/Hyperactivity Disorder: a population-based study of children aged 10 to 11 years. Psychosom Med. 2020;82(3):305–15.

8. Łojko D, Stelmach-Mardas M, Suwalska A. Diet quality and eating patterns in euthymic bipolar patients. Eur Rev Med Pharmacol Sci. 2019;23(3):1221–38.

9. Alkerwi A, Baydarlioglu B, Sauvageot N, Stranges S, Lemmens P, Shivappa N, et al. Smoking status is inversely associated with overall diet quality: Findings from the ORISCAV-LUX study. Clin Nutr. 2017;36(5):1275–82.

10. Steele CC, Pirkle JRA, Kirkpatrick K. Diet-induced impulsivity: Effects of a high-fat and a high-sugar diet on impulsive choice in rats. PLoS One. 2017;12(6):1–16.

11. Krishna S, Keralapurath MM, Lin Z, Wagner JJ, de La Serre CB, Harn DA, et al. Neurochemical and electrophysiological deficits in the ventral hippocampus and selective behavioral alterations caused by high-fat diet in female C57BL/6 mice. Neuroscience [Internet]. 2015;297:170–81. Available from: http://dx.doi.org/10.1016/j.neuroscience.2015.03.068

12. Sharma S, Fernandes MF, Fulton S. Adaptations in brain reward circuitry underlie palatable food cravings and anxiety induced by high-fat diet withdrawal. Int J Obes [Internet]. 2013;37(9):1183–91. Available from: http://dx.doi.org/10.1038/ijo.2012.197

13. Verlaet AAJ, Noriega DB, Hermans N, Savelkoul HFJ. Nutrition, immunological mechanisms and dietary immunomodulation in ADHD. Eur Child Adolesc Psychiatry. 2014;23(7):519–29.

14. Pelsser LMJ, Buitelaar JK, Savelkoul HFJ. ADHD as a (non) allergic hypersensitivity disorder: A hypothesis. Pediatr Allergy Immunol. 2009;20(2):107–12.

15. Wakimoto P, Block G. Dietary Intake, Dietary Patterns, and Changes With Age: An Epidemiological Perspective. Journals Gerontol Ser A Biol Sci Med Sci. 2001;56(Supplement 2):65–80.

16. Westenhoefer J. Age and gender dependent profile of food choice. Forum Nutr. 2005;57:44–51.

17. Schuit AJ, Van Loon AJM, Tijhuis M, Ocké MC. Clustering of lifestyle risk factors in a general adult population. Prev Med (Baltim). 2002;35(3):219–24.

18. Kinnunen MI, Suihko J, Hankonen N, Absetz P, Jallinoja P. Self-control is associated with physical activity and fitness among young males. Behav Med. 2012;38(3):83–9.

19. Cardol CK, Escamilla CI, Gebhardt WA, Perales JC. Exploring the direct or inverse association of physical activity with behavioral addictions and other self-regulation problems. Adicciones. 2019;31(1):18–32.

20. Pokhrel P, Schmid S, Pagano I. Physical Activity and Use of Cigarettes and E-Cigarettes Among Young Adults. Am J Prev Med [Internet]. 2020;58(4):580–3. Available from: https://doi.org/10.1016/j.amepre.2019.10.015

21. Gapin JI, Labban JD, Etnier JL. The effects of physical activity on attention deficit hyperactivity disorder symptoms: The evidence. Prev Med (Baltim) [Internet]. 2011;52(SUPPL.):S70–4. Available from: http://dx.doi.org/10.1016/j.ypmed.2011.01.022

22. Bardo MT, Compton WM. Does physical activity protect against drug abuse vulnerability? Drug Alcohol Depend [Internet]. 2015;153:3–13. Available from: http://dx.doi.org/10.1016/j.drugalcdep.2015.05.037

23. Sudlow C, Gallacher J, Allen N, Beral V, Burton P, Danesh J, et al. UK Biobank: An Open Access Resource for Identifying the Causes of a Wide Range of Complex Diseases of Middle and Old Age. PLoS Med. 2015;12(3):1–10.

24. Davis KAS, Coleman JRI, Adams M, Allen N, Breen G, Cullen B, et al. Mental health in UK Biobank Revised. MedRxiv. 2019;

25. Cole JB, Florez JC, Hirschhorn JN. Comprehensive genomic analysis of dietary habits in UK Biobank identifies hundreds of genetic associations. Nat Commun [Internet]. 2020;11(1):1–11. Available from: http://dx.doi.org/10.1038/s41467-020-15193-0

26. Fraley C, Raftery AE, Scrucca L, Murphy TB, Fop T. Package ‘mclust .’ 2020. p. 1–169.

27. Hill WD, Davies NM, Ritchie SJ, Skene NG, Bryois J, Bell S, et al. Genome-wide analysis identifies molecular systems and 149 genetic loci associated with income. Nat Commun [Internet]. 2019;10(1):1–16. Available from: http://dx.doi.org/10.1038/s41467-019-13585-5

28. van Buuren S, Groothuis-Oudshoorn K. mice: Multivariate imputation by chained equations in R. J Stat Softw. 2011;45(3):1–67.

29. Nieminen P, Lehtiniemi H, Vähäkangas K, Huusko A, Rautio A. Standardised regression coefficient as an effect size index in summarising findings in epidemiological studies. Epidemiol Biostat Public Heal. 2013;10(4):1–15.

30. Alamy M, Bengelloun WA. Malnutrition and brain development: An analysis of the effects of inadequate diet during different stages of life in rat. Neurosci Biobehav Rev [Internet]. 2012;36(6):1463–80. Available from: http://dx.doi.org/10.1016/j.neubiorev.2012.03.009

31. Tylee DS, Sun J, Hess JL, Tahir MA, Sharma E, Malik R, et al. Genetic correlations among psychiatric and immune-related phenotypes based on genome-wide association data. Am J Med Genet Part B Neuropsychiatr Genet. 2018;177(7):641–57.

32. Mills JS, Weinheimer L, Polivy J, Herman CP. Are there different types of dieters? A review of personality and dietary restraint. Appetite [Internet]. 2018;125:380–400. Available from: https://doi.org/10.1016/j.appet.2018.02.014

33. Sariyska R, Markett S, Lachmann B, Montag C. What Does Our Personality Say About Our Dietary Choices? Insights on the Associations Between Dietary Habits, Primary Emotional Systems and the Dark Triad of Personality. Front Psychol. 2019;10(November):1–11.

34. Pfeiler TM, Egloff B. Personality and eating habits revisited: Associations between the big five, food choices, and Body Mass Index in a representative Australian sample. Appetite. 2020;149(May 2019).

35. Forestell CA, Nezlek JB. Vegetarianism, depression, and the five factor model of personality. Ecol Food Nutr [Internet]. 2018;57(3):246–59. Available from: https://doi.org/10.1080/03670244.2018.1455675

36. Keller C, Siegrist M. Does personality influence eating styles and food choices? Direct and indirect effects. Appetite [Internet]. 2015;84:128–38. Available from: http://dx.doi.org/10.1016/j.appet.2014.10.003

37. Pfeiler TM, Egloff B. Examining the “Veggie” personality: Results from a representative German sample. Appetite [Internet]. 2018;120:246–55. Available from: https://doi.org/10.1016/j.appet.2017.09.005

38. Cooper RE, Tye C, Kuntsi J, Vassos E, Asherson P. The effect of omega-3 polyunsaturated fatty acid supplementation on emotional dysregulation, oppositional behaviour and conduct problems in ADHD: A systematic review and meta-analysis. J Affect Disord. 2016;190:474–82.

39. Swain DP, Franklin BA. Comparison of cardioprotective benefits of vigorous versus moderate intensity aerobic exercise. Am J Cardiol. 2006;97(1):141–7.

40. Byambasukh O, Snieder H, Corpeleijn E. Relation Between Leisure Time, Commuting, and Occupational Physical Activity With Blood Pressure in 125 402 Adults: The Lifelines Cohort. J Am Heart Assoc. 2020;9(4).

