## Supplemental Materials for "Diet, physical activity and behavioural disinhibition in middle-aged and older adults: a UK Biobank study"

belonging to publication:

Table of contents:


| Section | Description | | Page |
|---|---|---|---|
| **1** | **PCA to derive behavioural disinhibition scores** | | **2** |
| | ST1.1: | Selected UK Biobank items related to disinhibition, impulsivity, compulsivity and/or emotional instability and their binarised response categories | 3 |
| | ST1.2: | ICD-10 diagnoses related to impulsivity, compulsivity and/or emotional instability | 5 |
| | ST1.3: | Tetrachoric correlations between the nine disinhibited behaviour groups | 8 |
| | ST1.4: | PCA results extracting one, two and three principal components | 9 |
| **2** | **PCA to derive dietary components (DC1-4) and dietary groups (DC1-5)** | | **10** |
| | ST2.1 | UK Biobank items related to diet. Note that items related to alcohol intake are not included, since heavy drinking contributed to our outcome of interest | 11 |
| | ST2.2 | Restructured items included in the PCA and their factor loadings | 13 |
| | ST2.3 | Table S2.3. Correlations between DC1-4 | 15 |
| **3** | **Latent profile analysis to derive multivariate dietary groups (DC1-5)** | | **16** |
| | SF3.1 | Light's generalisation of Cohen's kappa (100 permutations, 2-16 PCs) | 16 |
| | SF3.2 | Increase in BIC upon adding latent participant clusters | 17 |
| **4** | **Outcomes when including age-by-predictor interaction terms** | | **18** |
| | ST4.1 | Regression estimates (β, SE) as reported in the main paper, after including an age-by-predictor interaction term | 19 |
| | ST4.2 | Regression estimates (β, SE) for age-by-predictor interaction terms | 20 |
| | ST4.3 | Simple slopes in age-deciles for the association between disinhibition diet | 21 |


PREPRINT

### Section 1: PCA to derive behavioural disinhibition scores

We performed a principal component analysis (PCA) on all disinhibition-related items. First, all datafields related to disinhibition, impulsivity, compulsivity and/or emotional instability were selected by two independent authors (Supplementary tables ST1.1-1.2). Non-binary items were binarized. Next, to ascertain a balanced representation of disinhibited behaviour types, cases were defined for nine groups of disinhibited behaviours (i.e. externalising, obsessive-compulsive, addiction, self-harm, personality, mania, risk-taking, cannabis use, smoking). Case-ness was defined such that it captured relatively extreme behaviours, aiming at a 5-10% endorsement rate. For instance, participants were considered a case for addiction when they either self-reported being or having been addicted, or had at any time received a diagnosis of substance dependence syndrome in hospital, resulting in 6.1% of the sample being a case.

Next, we performed a PCA based on tetrachoric correlations between behavioural groups (ST1.3). The theoretical framework underlying our approach is that impulsive, compulsive and emotionally unstable behaviours reflect a unitary disinhibited phenotype and share a common origin. Therefore, the single-component model was preferred a priori, and more complex models were only considered if the simpler model presented with statistical or interpretational shortcomings. All behaviours loaded positively on the first principal component, with factor loadings ranging from 0.335 to 0.708 (Table 2). Models with two and three principal components are shown in ST1.4. The two-PC model differentiates between behaviours related to substance abuse (PC2) and other disinhibited behaviours (PC1), but contains double loadings for self-harm and addiction. The three-PC model adds a component reflecting unstable personality/mania, but also has double loadings (addiction, personality) and triple loadings (self-harm). Hence, the single-component model was adopted. For each subject, a factor score was extracted with higher scores indicating a higher tendency for disinhibition. Finally, after trimming at 97.5% to replace extreme values by the cut-off value (i.e. 3.4), disinhibition scores ranged from -0.78 to 3.40 (M=-0.14, SD=0.93).

PRE-PRINT

Table S1.1. Selected UK Biobank items related to disinhibition, impulsivity, compulsivity and/or emotional instability and their binarised response categories.

| Data-field | Source | Description | Response categories (original) | Response categories (binary) |
|---|---|---|---|---|
| 1239 | IAV | Do you smoke tobacco now? | - No<br>- Yes, on all/most days<br>- Only occasionally | - No / only occasionally<br>- Yes, on all/most days |
| 1920 | IAV | Does your mood often go up and down? | - No<br>- Yes | - No<br>- Yes |
| 1940 | IAV | Are you an irritable person? | - No<br>- Yes | - No<br>- Yes |
| 1960 | IAV | Do you often feel fed-up? | - No<br>- Yes | - No<br>- Yes |
| 20544 | MHQ | Have you been diagnosed with one or more of the following mental health problems by a professional, even if you don't have it currently? (tick box) | - OCD<br>- Mania<br>- ADHD | - No<br>- Yes<br>(per diagnosis) |
| 20548 | MHQ | Try to remember a period when you were in a "high" or "irritable" state (tick box) (conditional to: ever had period of extreme irritability/excitability) | - More restless than usual<br>- Thoughts were racing<br>- Easily distracted<br>- More active than usual | - No<br>- Yes, ever had period of mania/irritability with at least one of these symptoms |
| 20401 | MHQ | Have you been addicted to or dependent on one or more things, including substances (not cigarettes/coffee) or behaviours (such as gambling)? | - No<br>- Yes | - No<br>- Yes |
| 20416 | MHQ | How often do you have six or more drinks on one occasion? (conditional to: frequency of drinking alcohol) | - Never<br>- Less than monthly<br>- Monthly<br>- Weekly<br>- (Almost) daily | - Never / less than weekly<br>- Weekly / (almost) daily |
| 20453 | MHQ | Have you taken cannabis | - No | - No / less than 11 times |

PRE-PRINT

| | | | | |
|---|---|---|---|---|
| | | (marijuana, grass etc.), even if it was a long time ago? | - Yes, 1-2 times<br>- Yes, 3-10 times<br>- Yes, 11-100 times<br>- Yes, more than 100 times | - Yes, 11 times / more |
| 20480 | MHQ | Have you deliberately harmed yourself, whether or not you meant to end your life? | - No<br>- Yes | - No<br>- Yes |
| 41202 and 41204 | LHR | Main or secondary ICD-10 diagnoses from hospital inpatient records | See supplementary table ST1.2 | - No<br>- Yes<br>(per diagnostic group) |

Abbreviations: IAV= initial assessment visit, MHQ= mental health questionnaire, LHR= linked health records, ICD-10= international classification of diagnoses, 10th edition.

PRE-PRINT

Table ST1.2. ICD-10 diagnoses related to impulsivity, compulsivity and/or emotional instability per diagnostic group.

| | | |
|---|---|---|
| Addiction | F102 | Dependence syndrome of alcohol |
| | F112 | Dependence syndrome of opioids |
| | F122 | Dependence syndrome of cannabinoids |
| | F132 | Dependence syndrome of sedatives or hypnotics |
| | F142 | Dependence syndrome of cocaine |
| | F152 | Dependence syndrome of other stimulants |
| | F162 | Dependence syndrome of hallucinogens |
| | F192 | Dependence syndrome of multiple drug use and use of other psychoactive substances |
| Externalising | F900 | Disturbance of activity and attention |
| | F901 | Hyperkinetic conduct disorder |
| | F908 | Other hyperkinetic disorders |
| | F909 | Hyperkinetic disorder unspecified |
| | F910 | Conduct disorder confined to the family context |
| | F911 | Unsocialised conduct disorder |
| | F912 | Socialised conduct disorder |
| | F913 | Oppositional defiant disorder |
| | F918 | Other conduct disorders |
| | F919 | Conduct disorder unspecified |
| | F920 | Depressive conduct disorder |
| | F928 | Other mixed disorders of conduct and emotions |
| | F929 | Mixed disorder of conduct and emotions unspecified |
| Mania | F300 | Hypomania |
| | F301 | Mania without psychotic symptoms |
| | F302 | Mania with psychotic symptoms |
| | F308 | Other manic episodes |
| | F309 | Manic episode unspecified |
| | F310 | Bipolar affective disorder current episode hypomanic |
| | F311 | Bipolar affective disorder current episode manic without psychotic symptoms |
| | F312 | Bipolar affective disorder current episode manic with psychotic symptoms |
| | F313 | Bipolar affective disorder current episode mild or moderate depression |
| | F314 | Bipolar affective disorder current episode severe depression w/o psychotic symptoms |
| | F315 | Bipolar affective disorder current episode severe depression with psychotic symptoms |

PRE-PRINT

| | | |
|---|---|---|
| | F316 | Bipolar affective disorder current episode mixed |
| | F317 | Bipolar affective disorder currently in remission |
| | F318 | Other bipolar affective disorders |
| | F319 | Bipolar affective disorder unspecified |
| OCD | F420 | Predominantly obsessional thoughts or ruminations |
| | F421 | Predominantly compulsive acts [obsessional rituals] |
| | F422 | Mixed obsessional thoughts and acts |
| | F428 | Other obsessive-compulsive disorders |
| | F429 | Obsessive-compulsive disorder unspecified |
| | F633 | Trichotillomania |
| | F950 | Transient tic disorder |
| | F951 | Chronic motor or vocal tic disorder |
| | F952 | Combined vocal and multiple motor tic disorder [de la Tourette] |
| | F958 | Other tic disorders |
| | F959 | Tic disorder unspecified |
| | F605 | Anankastic personality disorder |
| Personality | F603 | Emotionally unstable personality disorder |
| Risk-taking | F100 | Acute intoxication with alcohol |
| | F101 | Harmful use of acohol |
| | F110 | Acute intoxication of opioids |
| | F111 | Harmful use of opioids |
| | F120 | Acute intoxication cannabinoids |
| | F121 | Harmful use cannabinoids |
| | F130 | Acute intoxication of sedatives or hypnotics |
| | F131 | Harmful use of sedatives or hypnotics |
| | F140 | Acute intoxication of cocaine |
| | F141 | Harmful use of cocaine |
| | F150 | Acute intoxicationof other stimulants |
| | F151 | Harmful use of other stimulants |
| | F160 | Acute intoxication of hallucinogens |
| | F161 | Harmful use of hallucinogens |
| | F190 | Acute intoxication due to multiple drug use and use of other psychoactive substances |
| | F191 | Harmful use due to multiple drug use and use of other psychoactive substances |

PRE-PRINT

| | |
|---|---|
| F630 | Pathological gambling |
| F631 | Pathological fire-setting [pyromania] |
| F632 | Pathological stealing [kleptomania] |
| F638 | Other habit and impulse disorders |
| F639 | Habit and impulse disorder unspecified |

PRE-PRINT

Table S1.3. Tetrachoric correlations between the nine disinhibited behaviour groups.

| | OCD | Mania | Externalising | Personality | Risk-taking | Smoking | Addiction | Self-harm |
|---|---|---|---|---|---|---|---|---|
| Mania | 0.285 | | | | | | | |
| Externalising | 0.527 | 0.298 | | | | | | |
| Personality | 0.276 | 0.413 | 0.214 | | | | | |
| Risk-taking | 0.000 | 0.090 | 0.048 | 0.098 | | | | |
| Smoking | 0.096 | 0.156 | 0.065 | 0.119 | 0.196 | | | |
| Addiction | 0.278 | 0.295 | 0.287 | 0.221 | 0.298 | 0.357 | | |
| Self-harm | 0.318 | 0.322 | 0.239 | 0.265 | 0.075 | 0.217 | 0.366 | |
| Cannabis | 0.090 | 0.156 | 0.205 | 0.118 | 0.273 | 0.354 | 0.392 | 0.288 |

PRE-PRINT

Table S1.4. PCA results extracting one, two and three principal components. The two-PC model differentiates between behaviours related to substance abuse (PC2) and other disinhibited behaviours (PC1), but contains double loadings for self-harm and addiction. The three-PC model adds a component reflecting unstable personality/mania, but also has double loadings (addiction, personality) and triple loadings (self-harm).

| | Model: 1 PC | Model: 2 PCs | | Model: 3 PCs | | |
|---|---|---|---|---|---|---|
| | PC1 | PC1 | PC2 | PC1 | PC2 | PC3 |
| OCD | 0.591 | 0.816 | -0.153 | 0.847 | | |
| Mania | 0.613 | 0.634 | | | | 0.788 |
| Externalising | 0.589 | 0.739 | | 0.883 | | -0.113 |
| Personality | 0.536 | 0.692 | | -0.108 | -0.103 | 0.925 |
| Risk-taking | 0.335 | -0.223 | 0.686 | -0.287 | 0.658 | |
| Smoking | 0.475 | | 0.715 | -0.107 | 0.712 | |
| Addiction | 0.708 | 0.259 | 0.627 | 0.218 | 0.650 | |
| Self-harm | 0.637 | 0.482 | 0.283 | 0.272 | 0.278 | 0.295 |
| Cannabis | 0.552 | | 0.749 | | 0.788 | -0.167 |

PRE-PRINT

**Section 2: PCA to derive dietary components (DC1-4) and dietary groups (DC1-5)**

After removal of items related to alcohol intake, raw food frequency questionnaire (FFQ) data included nine continuous variables (e.g. tablespoons of vegetables), nine ordinal variables (e.g. oily fish intake frequency), five food type descriptions (e.g. bread type: brown/white/wholemeal/other), and one tick-box item assessing elimination of specific food groups (eggs, dairy, wheat, sugar; ST2.1). Ordinal intake frequency items were mapped to a continuous scale in days per month. Added salt (never, sometimes, often, always) was mapped to 0/0.333/0.667/1. Binary contrasts were created for each food type variable, such that each response category contrasted with all other categories (e.g. 'brown bread vs. any other type of bread') and that the never category contrasted with all other categories (e.g. 'I never eat bread vs. any type of bread'). One additional ordinal variable was created for fat content of milk ('no milk', 'skimmed milk', 'semi-skimmed milk', 'full-cream milk'). For the elimination item, we contrasted each restriction to having no restrictions (e.g. 'I never eat eggs vs. I do eat eggs, dairy, wheat and sugar') and to having no restrictions or other restrictions (e.g. 'I never eat eggs vs. no eggs restrictions'). Contrasts 'I never eat sugar vs. no restrictions' and 'I never eat sugar vs no sugar restrictions' were collinear ($r>0.85$), hence the latter was removed. The remaining 58 dietary items and contrasts are listed in ST2.2. All dietary items and contrasts were conditioned to age and sex, and continuous items were normalised applying rank-based inverted normal transformation.

Next, we performed a PCA with promax rotations, starting with one principal component and adding components one by one. Added components were retained when they contributed unique information, were interpretable and plausible, and remained stable upon including additional components to the model. The optimal model contained four dietary components (DC1-4) that were modestly correlated among each other (ST2.3).

Table S2.1. UK Biobank items related to diet. Note that items related to alcohol intake are not included, since heavy drinking contributed to our outcome of interest.

| Type | UKB ID | Item | Unit |
|---|---|---|---|
| Continuous | 1289 | Cooked vegetables | Tablespoons / day |
| | 1299 | Raw vegetables | Tablespoons / day |
| | 1309 | Fresh fruit | Pieces / day |
| | 1319 | Dried fruit | Pieces / day |
| | 1438 | Bread | Slices / week |
| | 1458 | Cereal | Bowls / week |
| | 1488 | Tea | Cups / day |
| | 1498 | Coffee | Cups / day |
| | 1528 | Water | Glasses / day |
| Frequency | 1329 | Oily fish | Times / week |
| | 1339 | Non-oily fish | Times / week |
| | 1349 | Processed meat | Times / week |
| | 1359 | Poultry | Times / week |
| | 1369 | Beef | Times / week |
| | 1379 | Lamb | Times / week |
| | 1389 | Pork | Times / week |
| | 1408 | Cheese | Times / week |
| | 1478 | Added salt | Never or rarely / sometimes / usually / always |
| Type | 1418 | Milk | Full-cream / semi-skimmed / skimmed / soy / other non-dairy / never |
| | 1428, 2654 | Spreads | Butter / block margarine / tub margarine / benecol / olive-oil based / sunflower-based / low-fat / other / never |
| | 1468 | Cereal | Bran / biscuit / oat / muesli / refined sugar-sweetened / never |
| | 1508 | Coffee | Decaffeinated / instant / ground / other / never |

PRE-PRINT

| | | | |
|---|---|---|---|
| | 1448 | Bread | Brown / white / wholegrain / other / never |
| Elimination | 6144 | I never eat (tick box) | Eggs / dairy / wheat / sugar |

PRE-PRINT

Table S2.2. Restructured items included in the PCA, and their factor loadings in the final model with four principal components (DC1-4)

| | **% endorsed** | **DC1** | **DC2** | **DC3** | **DC4** |
|---|---|---|---|---|---|
| Cooked vegetables | NA | 0.38 | 0.11 | 0.16 | -0.05 |
| Raw vegetables / salad | NA | 0.45 | 0.09 | 0.07 | -0.04 |
| Fresh fruit | NA | 0.45 | 0.04 | -0.04 | -0.09 |
| Dried fruit | NA | 0.48 | 0.00 | -0.05 | 0.07 |
| Bread | NA | -0.12 | -0.26 | -0.03 | 0.05 |
| Cereal | NA | 0.31 | -0.20 | -0.09 | 0.07 |
| Tea | NA | 0.03 | -0.05 | 0.01 | 0.04 |
| Coffee | NA | 0.00 | -0.17 | 0.19 | -0.09 |
| Water | NA | 0.34 | 0.15 | 0.02 | -0.01 |
| Oily fish | NA | 0.47 | 0.07 | 0.27 | -0.09 |
| Non-oily fish | NA | 0.33 | 0.02 | 0.33 | -0.11 |
| Processed meat | NA | -0.24 | -0.07 | 0.48 | 0.05 |
| Poultry | NA | -0.07 | 0.01 | 0.40 | -0.11 |
| Beef | NA | -0.05 | 0.00 | 0.73 | 0.02 |
| Lamb | NA | 0.09 | 0.04 | 0.75 | 0.05 |
| Pork | NA | 0.01 | 0.00 | 0.73 | 0.01 |
| Cheese | NA | 0.02 | -0.05 | 0.01 | 0.25 |
| Added salt | NA | -0.09 | 0.04 | 0.31 | 0.12 |
| Fat content of milk | NA | -0.06 | -0.01 | 0.05 | 0.83 |
| Full cream milk vs. any other milk | 6.0 | -0.12 | 0.08 | 0.03 | 0.31 |
| Semi-skimmed milk vs. any other milk | 66.2 | 0.01 | -0.11 | 0.06 | 0.55 |
| Skimmed milk vs. any other milk | 21.9 | 0.00 | -0.11 | 0.01 | -0.84 |
| Soy milk vs. any other milk | 4.7 | 0.11 | 0.28 | -0.18 | 0.07 |
| Other non-dairy milk vs. any other milk | 1.2 | 0.00 | 0.20 | -0.03 | 0.02 |
| Milk vs. 'I never drink milk' | 96.2 | 0.08 | -0.24 | -0.02 | 0.26 |
| Spread vs. 'I never use spreads' | 88.3 | -0.16 | -0.17 | 0.02 | 0.18 |
| Butter vs. any other spread | 41.8 | 0.05 | 0.09 | 0.15 | 0.29 |
| Block margarine vs. any other spread | <0.1 | -0.02 | 0.02 | 0.02 | 0.02 |
| Tub margarine vs. any other spread | 5.1 | -0.17 | 0.01 | -0.02 | -0.01 |
| Benecol vs. any other spread | 10.1 | 0.06 | -0.01 | -0.01 | -0.16 |

PRE-PRINT

| | | | | | |
|---|---|---|---|---|---|
| Olive-oil based spread vs. any other spread | 16.2 | 0.11 | -0.06 | -0.05 | -0.05 |
| Sunflower-based spread vs. any other spread | 19.3 | -0.07 | -0.09 | -0.06 | -0.14 |
| Low-fat spread vs. any other spread | 5.0 | -0.05 | -0.03 | -0.03 | -0.12 |
| Other spread vs. any other spread | 2.4 | -0.04 | 0.15 | -0.07 | 0.04 |
| White bread vs. any other bread | 20.6 | -0.55 | 0.09 | 0.05 | 0.01 |
| Brown bread vs. any other bread | 10.1 | -0.13 | 0.03 | 0.02 | 0.01 |
| Wholegrain bread vs. any other bread | 64.8 | 0.58 | -0.19 | -0.06 | -0.02 |
| Other bread vs. any other bread | 4.5 | -0.07 | 0.22 | 0.01 | 0.02 |
| Bread vs. 'I never eat bread' | 96.4 | -0.04 | -0.38 | -0.02 | 0.05 |
| Bran vs. any other cereal | 16.8 | 0.02 | -0.08 | 0.01 | -0.09 |
| Biscuit vs. any other cereal | 15.4 | -0.12 | -0.07 | 0.01 | -0.03 |
| Oat vs. any other cereal | 26.2 | 0.10 | 0.07 | 0.03 | -0.08 |
| Muesli vs. any other cereal | 25.5 | 0.26 | -0.02 | -0.06 | 0.17 |
| Refined sugar-sweetened cereal products | 16.0 | -0.33 | 0.09 | 0.02 | 0.02 |
| Cereal vs. 'I never eat cereal' | 84.2 | 0.28 | -0.23 | -0.09 | 0.10 |
| Decaffeinated coffee vs. any other coffee | 18.5 | 0.03 | -0.01 | -0.03 | -0.12 |
| Instant coffee vs. any other coffee | 51.2 | -0.31 | -0.08 | 0.02 | -0.09 |
| Ground coffee vs. any other coffee | 28.9 | 0.32 | 0.10 | 0.00 | 0.20 |
| Other coffee vs. any other coffee | 1.3 | -0.02 | 0.02 | -0.03 | -0.01 |
| Coffee vs. 'I never drink coffee' | 80.6 | 0.10 | -0.20 | 0.14 | -0.04 |
| 'I never eat eggs' vs. no restrictions | 2.5 | -0.07 | 0.50 | -0.06 | 0.04 |
| 'I never eat eggs' vs. no eggs restrictions | 10.1 | -0.07 | 0.24 | -0.05 | 0.05 |
| 'I never eat dairy' vs. no restrictions | 2.3 | -0.03 | 0.64 | -0.03 | 0.04 |
| 'I never eat dairy' vs. no dairy restrictions | 9.4 | -0.04 | 0.34 | -0.02 | 0.06 |
| 'I never eat wheat' vs. no restrictions | 2.6 | -0.06 | 0.64 | 0.07 | 0.05 |
| 'I never eat wheat' vs. no wheat restrictions | 10.6 | -0.06 | 0.34 | 0.05 | 0.07 |
| 'I never eat sugar' vs. no restrictions | 16.5 | 0.08 | -0.31 | 0.01 | -0.07 |
| I never eat eggs, sugar, wheat or dairy' vs. no restrictions | 19.9 | 0.06 | 0.51 | 0.03 | -0.09 |

PRE-PRINT

Table S2.3. Correlations between DC1-4

| | DC1 | DC2 | DC3 | DC4 |
|---|---|---|---|---|
| DC1: Prudent diet | 1.000 | | | |
| DC2: Wheat/dairy/egg elimination | 0.112 | 1.000 | | |
| DC3: Meat | -0.121 | -0.097 | 1.000 | |
| DC4: Full-cream dairy | -0.107 | -0.124 | 0.045 | 1.000 |

PRE-PRINT

**Section 3: Latent profile analysis to derive multivariate dietary groups (DC1-5)**

R-package 'mclust' [26] performs model-based clustering of participants based on parameterized finite Gaussian mixture models. Input to the clustering algorithm were dietary components DC1-4. We defined no a priori model constraints, and allowed a maximum of 16 clusters or subject groupings. Clustering is an iterative process, hence we performed 100 permutations and calculated Light's generalisation of Cohen's kappa's as an indicator of model stability, setting kappa>0.8 as our threshold for acceptable stability. Model fit was assessed using Bayesian information criterion (BIC), striving for maximal parsimony without compromising model fit. In addition, each cluster should represent no less than 1% of the full sample, and the model solution should be both interpretable and plausible. Figures S3.1 and S3.2 show that only the five-cluster solution has both stable model fit and acceptable model stability. Once the optimal model had been identified, group membership of each participant was set to the mode of classifications across 1,000 permuted model fittings.

Figure S3.1. Light's generalisation of Cohen's kappa's across one hundred permutations for models containing two to sixteen principal components. Kappa illustrates reproducibility of the model among permuted datasets and should ideally by 0.8 or higher.

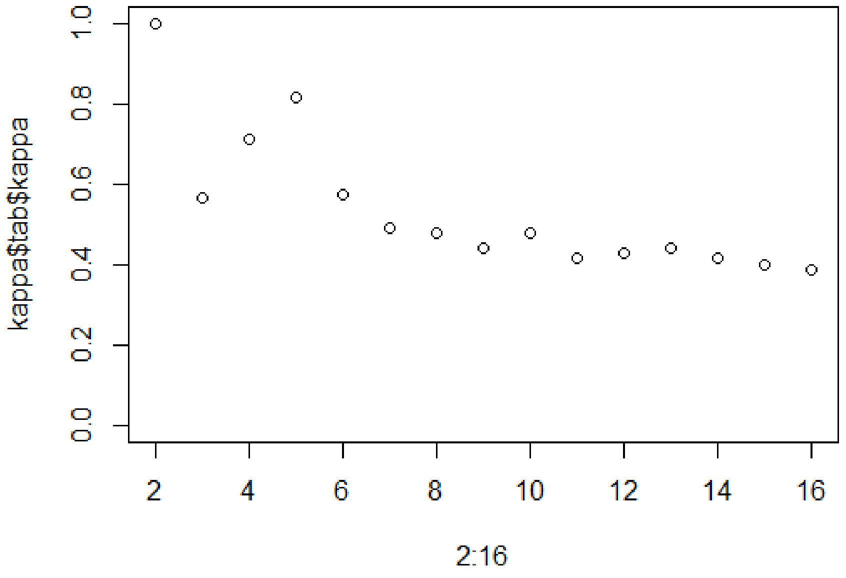

Figure S3.2. Increase in BIC upon adding latent participant clusters. Note that stabilisation starts after adding the fourth cluster, and is completed after adding the sixteenth cluster. VVV indicates a model with no *a priori* constraints.

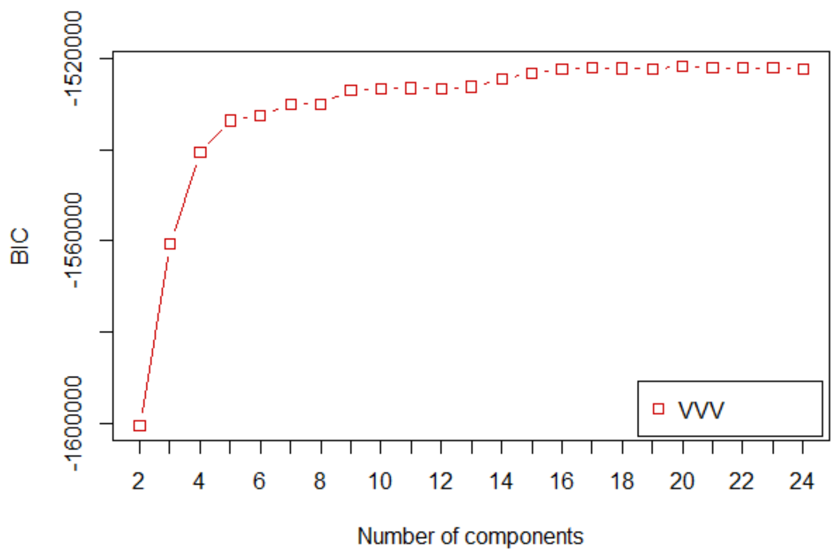


PRE-PRINT

**Section 4: Outcomes when including age-by-predictor interaction terms**

None of the associations reported in the main paper changed upon adding age-interaction effects (ST4.1). Moreover, most age-interaction effects were non-significant (ST4.2). In women, age-interactions with the unhealthy-vs-moderate-and-prudent, restricted-vs-moderate-and-prudent and low-fat dairy-vs-moderate-and-prudent group contrasts reached significance. In men, the age-by-DC2 interaction reached significance. For these interactions, we calculated the simple slopes of the dietary predictor for ten age-deciles (ST4.3). Note that a) the direction of effect remains the same across the age range, and b) the effect remains significant across the age range except at very late age. We conclude that some associations between diet and disinhibition may be stronger at younger age, especially in women, but that overall associations between diet and behavioural disinhibition are not moderated by age.

PRE-PRINT

Table S4.1. Associations as reported in the main paper, after including an age-by-predictor interaction term

| | | Men | | | Women | | |
|---|---|---|---|---|---|---|---|
| | | **β** | **SE** | **P-value** | **β** | **SE** | **P-value** |
| Single-predictor | DC1 | -0.036 | 0.004 | <0.0001 | -0.043 | 0.003 | <0.0001 |
| | DC2 | 0.031 | 0.004 | <0.0001 | 0.038 | 0.003 | <0.0001 |
| | DC3 | 0.041 | 0.004 | <0.0001 | -0.016 | 0.003 | <0.0001 |
| | DC4 | 0.023 | 0.004 | <0.0001 | 0.010 | 0.003 | 0.0036 |
| | Unhealthy-vs-moderate-and-prudent | 0.086 | 0.010 | <0.0001 | 0.096 | 0.009 | <0.0001 |
| | Restricted-vs-moderate-and-prudent | 0.125 | 0.016 | <0.0001 | 0.158 | 0.013 | <0.0001 |
| | Avoid meat-vs-moderate-and-prudent | 0.075 | 0.011 | <0.0001 | 0.153 | 0.011 | <0.0001 |
| | Low-fat dairy-vs-moderate-and-prudent | 0.041 | 0.011 | 0.0002 | 0.054 | 0.009 | <0.0001 |
| | MVPA | -0.007 | 0.004 | 0.0523 | -0.009 | 0.003 | 0.0053 |
| Multiple predictor | DC1 | -0.031 | 0.004 | <0.0001 | -0.049 | 0.003 | <0.0001 |
| | DC2 | 0.040 | 0.004 | <0.0001 | 0.044 | 0.003 | <0.0001 |
| | DC3 | 0.039 | 0.004 | <0.0001 | -0.018 | 0.003 | <0.0001 |
| | DC4 | 0.022 | 0.004 | <0.0001 | 0.010 | 0.003 | 0.0017 |
| | Unhealthy-vs-moderate-and-prudent | 0.085 | 0.010 | <0.0001 | 0.094 | 0.009 | <0.0001 |
| | Restricted-vs-moderate-and-prudent | 0.125 | 0.016 | <0.0001 | 0.157 | 0.013 | <0.0001 |
| | Avoid meat-vs-moderate-and-prudent | 0.075 | 0.011 | <0.0001 | 0.153 | 0.011 | <0.0001 |
| | Low-fat dairy-vs-moderate-and-prudent | 0.041 | 0.011 | 0.0002 | 0.053 | 0.009 | <0.0001 |
| | MVPA | -0.004 | 0.004 | 0.2522 | -0.003 | 0.003 | 0.3733 |

* β>0.02 & p<0.0028.

PRE-PRINT

Table S4.2. Age-by-predictor interaction terms, added to the models as described in the main paper.

| | | Men | | | Women | | |
|---|---|---|---|---|---|---|---|
| | | **β** | **SE** | **P-value** | **β** | **SE** | **P-value** |
| Single-predictor | DC1-by-age | -0.002 | 0.004 | 0.5530 | 0.011 | 0.003 | 0.0007 |
| | DC2-by-age | -0.019 | 0.004 | <0.0001 | -0.014 | 0.003 | <0.0001 |
| | DC3-by-age | <0.001 | 0.004 | 0.9122 | 0.003 | 0.003 | 0.3356 |
| | DC4-by-age | -0.004 | 0.004 | 0.2709 | 0.004 | 0.003 | 0.2653 |
| | Unhealthy-vs-moderate-and-prudent-by-age | -0.014 | 0.010 | 0.1521 | -0.031 | 0.009 | 0.0003* |
| | Restricted-vs-moderate-and-prudent-by-age | -0.044 | 0.016 | 0.0045 | -0.065 | 0.013 | <0.0001* |
| | Avoid meat-vs-moderate-and-prudent-by-age | -0.032 | 0.011 | 0.0061 | -0.027 | 0.011 | 0.0135 |
| | Low-fat dairy-vs-moderate-and-prudent-by-age | -0.022 | 0.011 | 0.0466 | -0.031 | 0.009 | 0.0005* |
| | MVPA-by-age | -0.005 | 0.004 | 0.1422 | 0.001 | 0.003 | 0.6528 |
| Multiple predictor | DC1-by-age | 0.004 | 0.004 | 0.2564 | 0.013 | 0.003 | 0.0001 |
| | DC2-by-age | -0.021 | 0.004 | <0.0001* | -0.015 | 0.003 | <0.0001 |
| | DC3-by-age | -0.002 | 0.004 | 0.6387 | 0.002 | 0.003 | 0.4604 |
| | DC4-by-age | -0.007 | 0.004 | 0.0486 | 0.003 | 0.003 | 0.3806 |
| | Unhealthy-vs-moderate-and-prudent-by-age | -0.014 | 0.010 | 0.1364 | -0.030 | 0.009 | 0.0004* |
| | Restricted-vs-moderate-and-prudent-by-age | -0.045 | 0.016 | 0.0044 | -0.065 | 0.013 | <0.0001* |
| | Avoid meat-vs-moderate-and-prudent-by-age | -0.030 | 0.011 | 0.0061 | -0.027 | 0.011 | 0.0130 |
| | Low-fat dairy-vs-moderate-and-prudent-by-age | -0.023 | 0.011 | 0.0451 | -0.031 | 0.009 | 0.0006* |
| | MVPA-by-age | -0.007 | 0.004 | 0.0821 | -0.001 | 0.003 | 0.8340 |

* β>0.02 & p<0.0028.

PRE-PRINT

ST4.3 Simple slopes at ten age-deciles for the association between disinhibition and the dietary predictor, in those instances where the age-by-dietary predictor interaction term reached significance.

| | | | Single-predictor model | | | Multiple-predictor model | | |
|---|---|---|---|---|---|---|---|---|
| | | **Age (SD)** | **β** | **SE** | **P-value** | **β** | **SE** | **P-value** |
| Women | Unhealthy-vs-moderate-and-prudent | -2.0 | 0.151 | 0.019 | <0.0001 | 0.149 | 0.019 | <0.0001 |
| | | -1.6 | 0.138 | 0.016 | <0.0001 | 0.136 | 0.016 | <0.0001 |
| | | -1.1 | 0.124 | 0.013 | <0.0001 | 0.123 | 0.013 | <0.0001 |
| | | -0.7 | 0.111 | 0.010 | <0.0001 | 0.110 | 0.010 | <0.0001 |
| | | -0.3 | 0.098 | 0.009 | <0.0001 | 0.097 | 0.009 | <0.0001 |
| | | 0.2 | 0.085 | 0.009 | <0.0001 | 0.084 | 0.009 | <0.0001 |
| | | 0.6 | 0.072 | 0.010 | <0.0001 | 0.071 | 0.010 | <0.0001 |
| | | 1.0 | 0.059 | 0.012 | <0.0001 | 0.058 | 0.012 | <0.0001 |
| | | 1.5 | 0.046 | 0.015 | 0.0026 | 0.045 | 0.015 | 0.0031 |
| | | 1.9 | 0.032 | 0.018 | 0.0754 | 0.032 | 0.018 | 0.0823 |
| | Restricted-vs-moderate-and-prudent | -2.0 | 0.288 | 0.030 | <0.0001 | 0.288 | 0.030 | <0.0001 |
| | | -1.6 | 0.260 | 0.025 | <0.0001 | 0.260 | 0.025 | <0.0001 |
| | | -1.1 | 0.232 | 0.020 | <0.0001 | 0.232 | 0.020 | <0.0001 |
| | | -0.7 | 0.204 | 0.016 | <0.0001 | 0.204 | 0.016 | <0.0001 |
| | | -0.3 | 0.176 | 0.014 | <0.0001 | 0.175 | 0.014 | <0.0001 |
| | | 0.2 | 0.147 | 0.013 | <0.0001 | 0.147 | 0.013 | <0.0001 |
| | | 0.6 | 0.119 | 0.015 | <0.0001 | 0.119 | 0.015 | <0.0001 |
| | | 1.0 | 0.091 | 0.018 | <0.0001 | 0.091 | 0.018 | <0.0001 |
| | | 1.5 | 0.063 | 0.022 | 0.0049 | 0.063 | 0.022 | 0.0051 |
| | | 1.9 | 0.035 | 0.027 | 0.1996 | 0.035 | 0.027 | 0.2026 |
| | Low-fat dairy-vs-moderate-and-prudent | -2.0 | 0.111 | 0.019 | <0.0001 | 0.110 | 0.019 | <0.0001 |
| | | -1.6 | 0.098 | 0.016 | <0.0001 | 0.097 | 0.016 | <0.0001 |
| | | -1.1 | 0.085 | 0.013 | <0.0001 | 0.085 | 0.013 | <0.0001 |
| | | -0.7 | 0.072 | 0.011 | <0.0001 | 0.072 | 0.011 | <0.0001 |
| | | -0.3 | 0.059 | 0.009 | <0.0001 | 0.059 | 0.009 | <0.0001 |
| | | 0.2 | 0.047 | 0.009 | <0.0001 | 0.046 | 0.009 | <0.0001 |

PRE-PRINT

| | | | | | | | | |
|---|---|---|---|---|---|---|---|---|
| | | 0.6 | 0.034 | 0.010 | 0.0006 | 0.033 | 0.010 | 0.0007 |
| | | 1.0 | 0.021 | 0.012 | 0.0830 | 0.021 | 0.012 | 0.0898 |
| | | 1.5 | 0.008 | 0.015 | 0.5816 | 0.008 | 0.015 | 0.6027 |
| | | 1.9 | -0.005 | 0.018 | 0.8032 | -0.005 | 0.018 | 0.7833 |
| Men | DC2 | -2.4 | | | | 0.090 | 0.010 | <0.0001 |
| | | -1.9 | | | | 0.080 | 0.008 | <0.0001 |
| | | -1.4 | | | | 0.070 | 0.007 | <0.0001 |
| | | -0.9 | | | | 0.059 | 0.005 | <0.0001 |
| | | -0.4 | | | | 0.049 | 0.004 | <0.0001 |
| | | 0.0 | | | | 0.039 | 0.004 | <0.0001 |
| | | 0.5 | | | | 0.029 | 0.004 | 0.0006 |
| | | 1.0 | | | | 0.019 | 0.005 | 0.0003 |
| | | 1.5 | | | | 0.009 | 0.007 | 0.1965 |
| | | 2.0 | | | | -0.002 | 0.008 | 0.8416 |

PRE-PRINT
